# Predictive performance of seven clinical surrogates of visceral adipose tissue for cardiovascular mortality: A sub-analysis of 102,385 adults from the Mexico City Prospective Study

**DOI:** 10.64898/2026.03.02.26347453

**Authors:** Jesús Ernesto Martínez-Luna, María Fernanda Suárez-Velázquez, Mario Cesar Torres-Chávez, Guillermo C. Cardoso-Saldaña, Juan Reyes-Barrera, Jaime Berumen-Campos, Pablo Kuri-Morales, Roberto Tapia-Conyer, Jesus Alegre-Díaz, Carlos A. Fermín-Martínez, Jacqueline A. Seiglie, Omar Yaxmehen Bello-Chavolla, Neftali Eduardo Antonio-Villa

**Author notes:** **Correspondence:** 1. Neftali Eduardo Antonio-Villa. Department of Endocrinology, Instituto Nacional de Cardiología Ignacio Chávez, Juan Badiano 1, Belisario Domínguez Secc 16, Tlalpan, 14080, Mexico City, Mexico., 2. Omar Yaxmehen Bello-Chavolla. Division of Research, Instituto Nacional de Geriatría, Anillo Perif, 2767, San Jerónimo Lídice, La Magdalena Contreras, 10200, Mexico City, Mexico. These authors contributed equally to the drafting of this work (Jesús Ernesto Martínez-Luna, Neftali Eduardo Antonio-Villa). Joint Last authorship (Jacqueline A. Seiglie, Omar Yaxmehen Bello-Chavolla, Neftali Eduardo Antonio-Villa). **Disclosures:** Nothing to disclose.

## Abstract

**BACKGROUND:** Visceral adipose tissue (VAT) has been associated with cardiovascular disease (CVD) mortality. However, the comparative performance of VAT-related clinical surrogates remains poorly characterized.

**OBJECTIVES:** To evaluate the performance of seven VAT-related clinical surrogates for predicting CVD and cause-specific CVD mortality.

**METHODS:** We analyzed data from the Mexico City Prospective Cohort, a population-based prospective cohort study, with baseline recruitmetn between 1998 – 2004 and ongoing mortality follow-up. CVD mortality included deaths from cardiac, stroke-related, and other vascular causes. Seven VAT-related surrogates (METS-VF, CVAI, EVA, DAAT, LAAP, VAI, and DAI) were estimated using clinical, biochemical, and anthropometric data at baseline. Associations with outcomes were evaluated using Cox regression models to estimate adjusted hazard ratios (aHRs). Discrimination was assessed with Harrell’s C-statistic (Cs) and fixed-point at 10-years receiver operating characteristic (ROC) curves, and calibration with slope plots.

**RESULTS:** In a subsample of 102,385 participants (median age: 47 years; 67% female), 4,068 (3.97%) died from any CVD causes. METS-VF (Cs: 0.722; aHR: 1.17, 95% CI: 1.12–1.23), EVA (Cs: 0.72; 1.14, 1.12–1.23), CVAI (Cs: 0.70; 1.13, 1.09–1.18), and DAAT (Cs: 0.626; 1.13, 1.09– 1.18) were positively associated with CVD mortality and showed the highest predictive capacity among the surrogates. Adding METS-VF to a CVD risk score among individuals classified as intermediate risk improved discrimination for CVD mortality.

**CONCLUSIONS:** In this large cohort of Mexican adults, four VAT-related clinical surrogates, particularly METS-VF, demonstrated good discriminatory performance for long-term CVD mortality. These indices could help to identify individuals with high VAT accumulation and high CVD risk in resource-limited settings.

## INTRODUCTION

Cardiovascular disease (CVD) remains the leading cause of death globally, responsible for approximately 18 million deaths annually and a substantial share of premature mortality in low- and middle-income countries (LMICs) ^1,2^. Although CVD mortality rates have declined in Latin America, population growth and aging have increased the absolute number of deaths, and CVDs still accounted for 1.1 million deaths in 2021 ^3–5^. In Mexico, deaths attributable to CVD, including ischemic heart disease, cerebrovascular disease and hypertension-related deaths have consistently ranked among the top causes of mortality in national vital statistics ^6^. This trend could be explained by the rising prevalence of cardiometabolic comorbidities in the country, such as type 2 diabetes, hypertension, dyslipidemia, and obesity ^7^.

Obesity, particularly central adiposity, is closely linked to cardiometabolic risk through the deposition of ectopic fat, most notably the accumulation of visceral adipose tissue (VAT). VAT is a metabolically active depot characterized by enhanced lipolysis, pro-inflammatory adipokines, hepatic insulin resistance, and atherogenic dyslipidemia ^8–10^. Furthermore, this dysfunctional profile has been associated with hepatic steatosis, arterial stiffness, and accelerated atherosclerotic disease, ultimately contributing to increase the progression to any CVD events ^11–16^. Thus, VAT emerges as a key mechanistic link between obesity and CVD mortality. Hence, the assessment of VAT has recently been proposed as a strategy to measure cardiometabolic risk, the burden of obesity-related CVD and its residual cardiovascular risk ^17^.

Although traditionally used anthropometric measures, such as body mass index (BMI) and waist circumference (WC), have been widely used in population-based studies to assess the effect of obesity and its effect on adverse cardiometabolic outcomes, these metrics do not capture inter-individual variation in fat distribution. Although gold-standard methods to measure VAT, included computed tomography (CT), magnetic resonance imaging (MRI), and dual-energy X-ray absorptiometry (DXA), their routine implementation is impractical for population-level assessment due to cost, access, and radiation/logistical constraints ^18^. To improve the estimation of VAT, the International Atherosclerosis Society and the International Chair on Cardiometabolic Risk Working Group on Visceral Obesity have endorsed the use of clinical surrogates of VAT to identify individuals with excess VAT in clinical and epidemiological settings, going beyond traditional indices such as BMI and WC ^19^. These indices are accessible, simple, low-cost, and provide a scalable approach in primary-care to estimate VAT in LMICs. Although prior studies have validated diverse VAT surrogates for cardiometabolic outcomes, many are limited by cross-sectional designs, short follow-up, or reliance on intermediate endpoints ^20–22^. These gaps leave uncertainty about which clinical surrogate of VAT best identifies long-term risk of CVD death and whether its performance generalizes to Latin American populations with distinct adiposity phenotypes and with a high burden of cardiometabolic comorbidities ^23–26^.

Therefore, in this study, we aimed to evaluate the performance of seven VAT-related clinical surrogates for predicting CVD and CVD cause-specific mortality using data from Mexican adults enrolled in the Mexico City Prospective Study (MCPS).

## METHODS

### Data source

We analyzed data from participants enrolled in the MCPS, a prospective, population-based cohort study with a baseline survey conducted from 1998 to 2004. Details of participant recruitment and follow-up have been published elsewhere ^27^. Briefly, the MCPS enrolled adults aged > 35 years living in the municipalities of Coyoacán and Iztapalapa in Mexico City. In total, 159,517 individuals were recruited, and trained nurses collected data regarding sociodemographic, lifestyle, and health-related information through electronic questionnaires. Standardized measurements of height, weight, WC, hip circumference (HC), and sitting blood pressure (BP) were obtained using calibrated instruments. Additionally, a non-fasting 10 mL blood sample was collected from each participant for subsequent analysis. Blood samples were shipped to the University of Oxford for analysis and storage. Available plasma samples from the baseline examination were analyzed using nuclear magnetic resonance (NMR) spectroscopy at Nightingale Health Ltd (Kuopio, Finland) and the Clinical Trial Service Unit’s Wolfson Laboratory, quantifying over 249 plasma biomarkers. For our analyses, we excluded individuals with a self-reported history of CVD (n = 2,871, 1.8%), adults living with diabetes, defined as self-reported diagnosis, use of glucose-lowering medication, or HbA1c ≥ 6.5% in undiagnosed participants ^28^ (n = 29,948, 18.8%), individuals aged ≥ 80 years (n = 4,213, 2.6%), those with BMI < 18.5 kg/m² or ≥ 40 kg/m² (n = 3,801, 2.4%), and those with incomplete data to estimate our visceral fat indexes or with extreme biochemical values required for VAT indexes estimation, defined as values +/- 3 standard deviations from their mean (SD) (n = 8,558, 5.4%). The decision to exclude individual living with diabetes was based on previous evidence that show differences in VAT composition among these individuals ^29^. All participants provided written informed consent. The corresponding ethics committees at the Mexican Ministry of Health, the Mexican National Council for Science and Technology, and the University of Oxford approved the study protocol. All study participants provided written informed consent. This report adheres to STROBE guidelines for prospective cohort studies (**Supplementary Table 1**).

### Outcome definition

Our main analyses focused on two major outcomes: CVD mortality and CVD cause-specific mortality. CVD mortality was defined according to Bosco Elliot *et al.* using International Classification of Diseases 10th Revision (ICD-10) codes, as specified in **Supplementary Table 2** ^30^. Cause-specific CVD mortality was grouped into three main categories: cardiac-related mortality, stroke-related mortality, and other CVD causes of mortality. Participants who were alive or died from non-CVD causes were considered as censorships. Mortality follow-up of was done through electronic probabilistic linkage to Mexican death registries up to September 30th, 2022, and causes of death were classified by cohort personnel according to the ICD-10.

### Surrogates of visceral adipose tissue

We aimed to externally validate the performance of seven previously described VAT surrogates to predict our studied outcomes. The estimated VAT surrogates were: Metabolic score for visceral fat (METS-VF) ^31^, Estimate of visceral adipose tissue area (EVA) ^32^, Chinese visceral adiposity index (CVAI)^33^, Deep abdominal adipose tissue (DAAT)^34^, Lipid accumulation product (LAP)^35^, Visceral adiposity index (VAI)^36^ and Dysfunctional adiposity index (DAI)^37^. Each index uses anthropometric, biochemical and clinical parameters which were extracted and adapted to the MCPS baseline evaluation. The formulae for each index are provided in **Supplementary Table 3**. Given that each index has a different scoring scale, for the association, discrimination, and calibration analyses, we standardized all indices to reflect increases of 1 standard deviation (1-SD) from the mean to enable direct comparison.

### Variable Assessment

For this analysis, we extracted key variables from the MCPS baseline dataset and grouped them into five domains: sociodemographic, lifestyle, clinical, anthropometric, and biochemical characteristics. Sociodemographic variables included age at enrollment, sex, municipality of residence (Coyoacán or Iztapalapa), and social development index (SDI) (ie, Índice de Desarrollo Social de la Ciudad de México por Manzana^38^) categorized as very low, low, intermediate, and high. Lifestyle variables included poor diet quality (defined as < 4 servings/week of fruits and vegetables) according to the composite previously applied to MCPS by Ferrero-Hernandez *et al.* ^39^, smoking status (never, former and current), alcohol consumption (never, former, > 3 times/month, > 2 times/week) and physical activity status (never, ≤ 2 times/week, ≥ 3/week). Clinical variables included arterial hypertension (self-report, antihypertensive medication use, or BP measurements ≥140/90 mmHg in 3 different readings ^40^), any dyslipidemia (defined as ≥1 of the following: triglycerides ≥ 150 mg/dL, total cholesterol ≥ 200 mg/dL, high density lipoprotein cholesterol HDL-C <50 mg/dL if was female and < 40 mg/dL if was male and low density lipoprotein cholesterol LDL-C ≥ 100 mg/dL ^41^), overweight (BMI ≥ 25 and < 30 kg/m^2^), obesity (BMI ≥ 30 kg/m2), metabolic syndrome (defined with ATP III criteria ^41^). Anthropometric variables included BMI, WC, and waist-to-height ratio (WtHR). Biochemical assessment included glucose (mg/dL), HbA1c (%), total cholesterol (mg/dL), HDL-C (mg/dL), LDL-C (mg/dL), and triglycerides (mg/dL).

### Statistical analysis

Participant characteristics are presented for the overall sample and stratified by CVD mortality status. To assess the association of each clinical surrogate with the evaluated outcomes, we fitted Cox proportional hazards regression models to estimate hazard ratios (HRs). Then, to diminish the effect of potential confounding, we adjusted these models for age, sex, poor diet quality, physical activity status, and metabolic syndrome, which were determined by creating a directed acyclic graph (DAG) following the recommendations of Tennant et al ^42^. The DAG used in these analyses are presented in **Supplementary Figure 1**. To evaluate the time-range discrimination performance of each clinical surrogate with our outcomes, we estimated Harrell’s C-statistic (C-statistic) for all indices based on unadjusted and adjusted Cox proportional hazards regression models. Then, fixed-point discrimination at 10 years was evaluated using the area under the receiver operating characteristic curve (AUROC). For the assessment of discrimination over the ∼20 years of follow-up, we performed time-dependent ROC curves, obtaining an integrated area under the curve (AUC) by the performance across follow-up years. For both measures, values close to 1 indicate good discrimination ability, whereas values close to 0.5 indicate poor discrimination. Calibration was evaluated by calculating the ratio of the observed 10-year survival (estimated using the Kaplan–Meier estimator) to the average predicted risk, where a ratio close to 1 indicates good agreement between predicted and observed event rates. These results were plotted as calibration curves, where the calibration slope should ideally be close to 1, for which values below or above 1 indicate underestimation or overestimation of risk, respectively. To determine meaningful thresholds, we identified the optimal cut-off points using maximization of the log-rank statistic, and they were presented in units of each surrogate. Finally, as a complementary analysis, we assessed the predictive value of our VAT surrogates combined with Globorisk, a validated model in the Mexican adult population aged ≥30 years that predicts the 10-year risk of CVD in healthy patients, compared with the performance of Globorisk alone, stratifying the participants according to the risk of mortality into: very low risk (<3%), low risk (3–6%), intermediate risk (7–9%), high risk (10–14%), and very high risk (≥15%) ^43^. For this, we performed Cox proportional hazards regression models to calculate the C-statistic and obtain Δ-Bayesian Information Criteria (BIC) and Δ-C-statistic for the overall sample and across all categories of risk. Higher Δ-C-statistic indicates a better improvement in the outcome prediction, while lower Δ-BIC represents better improvement in model adjustment. All analyses were performed using R Studio, and the code is publicly available in a GitHub repository and can be accessed at: https://github.com/neftalivilla/VAT_INDEXES_MCPS.

## RESULTS

### Baseline characteristics

Of the 159,517 participants originally enrolled, 57,132 (35.8%) were excluded, leaving a final analytic sample of 102,385 participants (**Figure 1**). The baseline characteristics stratified by outcome and CVD-related mortality are shown in **Table 1**. Briefly, most of the included sample were female (67%), overall median age of 47 years (IQR: 40–57) living in low SDI (43%). Regarding lifestyle characteristics, 47% consumed a poor-quality diet, 33% were active smoker, 53% consumed alcohol ≥2 times per week, and 77% reported no physical activity. Regarding clinical and anthropometric variables, 93% had any dyslipidemia (mostly driven by a prevalence of 85% low-HDL cholesterol), while the median BMI was 28.2 kg/m² (IQR: 25.5–31.2), classifying 80% of the sample as having either overweight or obesity. When stratified by CVD-related mortality, participants who died from any CVD tended to be older (66 years vs. 46 years), predominantly male (43% vs. 32%), and had a higher prevalence of hypertension (61% vs. 33%) and metabolic syndrome (52% vs. 37%). They also exhibited a higher WC (96 cm vs. 92 cm), although there were no differences in BMI. Among our estimated VAT indices, METS-VF, EVA, CVAI, and DAAT were higher in participants who died from CVD, while LAP, DAI, and VAI showed similar distributions across both groups. Sex-stratified characteristics are presented in **Supplementary Tables 4** and **Supplementary Table 5.**

**Figure 1:**
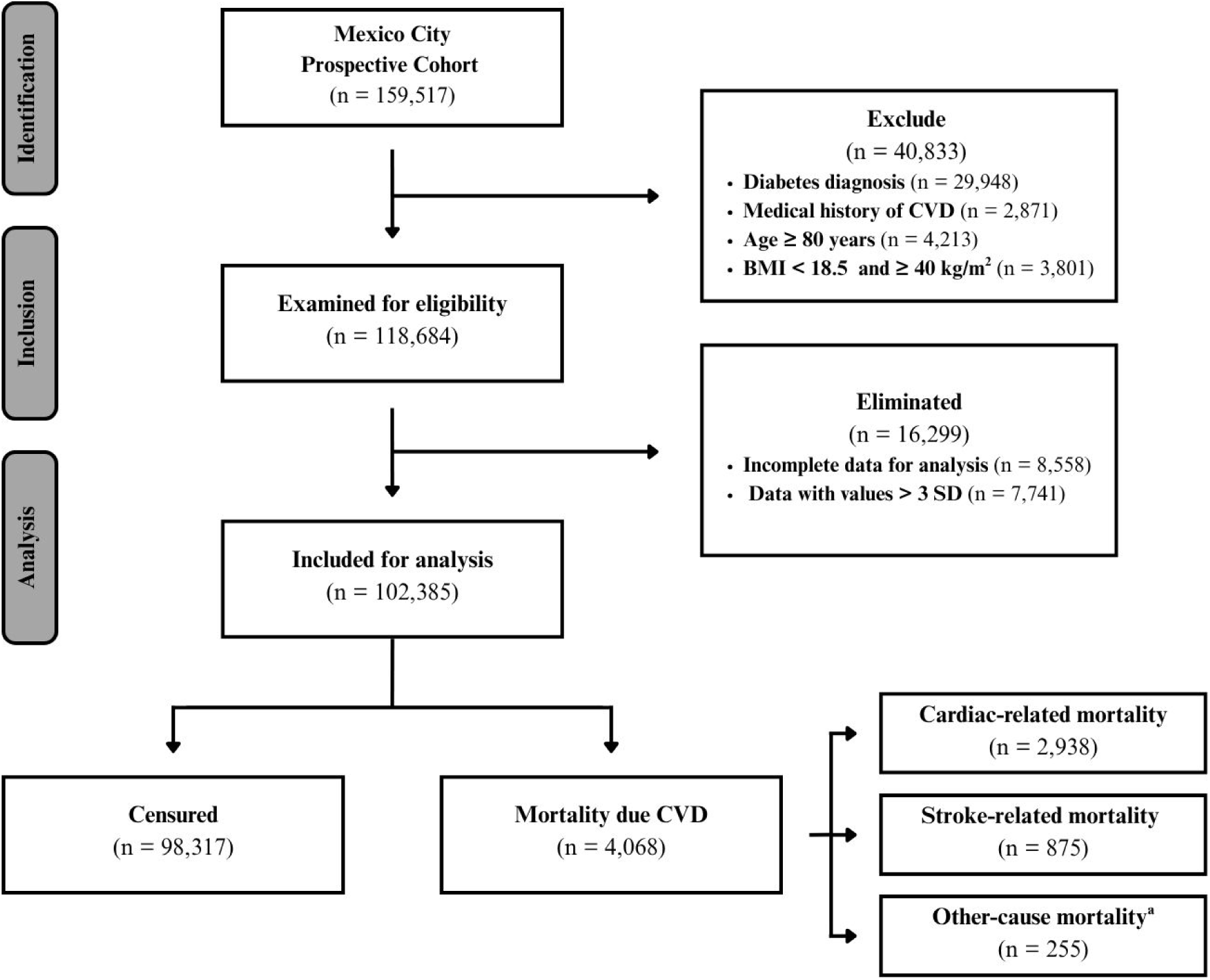
Flowchart of the participants from the Mexico City Prospective Cohort included in the analysis. Footnote: ^a^ Mortality due to CVD is not included in Cardiac-related and Stroke-related mortality Abbreviations: CVD, cardiovascular disease; BMI, body mass index; SD, standard deviation

**Table 1:** Baseline characteristics of participants included in analysis, stratified according to outcome.

| Characteristics | Overall sample<br>*n* = 102,385 | Censored<br>*n* = 98,317 | Cardiovascular-related<br>mortality<br>*n* = 4,068 |
| --- | --- | --- | --- |
| *Demographics* |  |  |  |
| Age, (years) | 47 (40, 57) | 46 (40, 56) | 66 (55, 73) |
| Female, (%) | 68,867 (67%) | 66,556 (68%) | 2,311 (57%) |
| Social development index, (%) |  |  |  |
| Very Low | 28,695 (29%) | 22,681 (29%) | 1,014 (26%) |
| Low | 41,688 (43%) | 39,925 (42%) | 1,763 (45%) |
| Intermediate | 15,030 (15%) | 14,318 (15%) | 712 (18%) |
| High | 12,498 (13%) | 12,037 (13%) | 461 (12%) |
| Unknown | 4,474 | 4,356 | 118 |
| County, (%) |  |  |  |
| Iztapalapa | 59,909 (59%) | 57,410 (58%) | 2,499 (61%) |
| Coyoacán | 42,476 (41%) | 40,907 (42%) | 1,569 (39%) |
| *Lifestyle* |  |  |  |
| Poor quality diet, (%) | 48,032 (47%) | 46,087 (47%) | 1,945 (48%) |
| Smoking, (%) |  |  |  |
| Never | 49,411 (48%) | 47,493 (48%) | 1,918 (47%) |
| Former | 19,067 (19%) | 18,021 (18%) | 1,046 (26%) |
| Current | 33,907 (33%) | 32,803 (33%) | 1,104 (27%) |
| Alcohol consumption, (%) |  |  |  |
| Never | 19,130 (19%) | 18,289 (19%) | 841 (21%) |
| Former | 6,350 (6.2%) | 6,151 (6.3%) | 199 (4.9%) |
| > 3 times per month | 23,023 (22%) | 21,890 (22%) | 1,133 (28%) |
| > 2 times per week | 53,882 (53%) | 51,987 (53%) | 1,895 (47%) |
| Physical activity, (%) |  |  |  |
| Never | 78,635 (77%) | 75,499 (77%) | 3,136 (77%) |
| ≤ 2 times per week | 8,640 (8.4%) | 8,363 (8.5%) | 277 (6.8%) |
| ≥ 3 times per week | 15,110 (15%) | 14,445 (15%) | 655 (16%) |
| *Clinical features* |  |  |  |
| Hypertension, (%) | 34,729 (34%) | 32,229 (33%) | 2,500 (61%) |
| Dyslipidemia, (%) | 94,883 (93%) | 91,186 (93%) | 3,697 (91%) |
| Low HDL-C (<40/50 mg/dL) | 87,317 (85%) | 83,929 (85%) | 3,388 (83%) |
| High LDL-C (≥130 mg/dL) | 3,049 (3.0%) | 2,934 (3.0%) | 115 (2.8%) |
| High total cholesterol (≥200 mg/dL) | 16,359 (16%) | 15,708 (16%) | 651 (16%) |
| High tryglicerides (≥150 mg/dL) | 30,540 (30%) | 29,385 (30%) | 1,155 (28%) |
| Overweight, (%) | 47,063 (46%) | 45,316 (46%) | 1,747 (43%) |
| Obesity, (%) | 34,841 (34%) | 33,335 (34%) | 1,506 (37%) |
| Metabolic syndrome, (%) | 38,374 (37%) | 36,248 (37%) | 2,126 (52%) |
| *Antropometric* |  |  |  |
| Body Mass Index, (kg/m2) | 28.2 (25.6, 31.2) | 28.2 (25.6, 31.2) | 28.5 (25.6, 31.7) |
| Waist circumference, (cm) | 93 (86, 99) | 92 (86, 99) | 96 (90, 103) |
| Waist to height ratio, (%) | 0.59 (0.55, 0.64) | 0.59 (0.55, 0.64) | 0.62 (0.57, 0.67) |
| *Biochemical assessment* |  |  |  |
| Glucose, (mg/dl) | 73 (62, 86) | 73 (62, 86) | 78 (65, 93) |
| HbA1c, (%) | 5.45 (5.17, 5.72) | 5.45 (5.17, 5.63) | 5.54 (5.35, 5.81) |
| Total cholesterol, (mg/dl) | 165 (141, 188) | 165(141, 188) | 164 (139, 188) |
| HDL-C, (mg/dl) | 39 (34, 43) | 38 (34, 43) | 38 (33, 43) |
| LDL-C, (mg/dl) | 65 (54, 77) | 65 (54, 77) | 64 (52, 77) |
| Triglycerides, (mg/dl) | 124 (96, 158) | 124 (96, 158) | 123 (96, 155) |
| *Surrogates evaluated* |  |  |  |
| METS-VF | 6.99 (6.67, 7.26) | 6.98 (6.66, 7.24) | 7.31 (7.06, 7.50) |
| CVAI | 121 (94, 149) | 120 (93, 147) | 150 (126, 174) |
| EVA | 114 (88, 143) | 113 (87, 141) | 146 (122, 171) |
| DAAT | 157 (119, 201) | 156 (118, 200) | 185 (148, 225) |
| LAP | 45 (31, 62) | 45 (31, 62) | 49 (34, 66) |
| DAI | 1.43 (1.09, 1.85) | 1.43 (1.09, 1.85) | 1.44 (1.11, 1.85) |
| VAI | 2.41 (1.81, 3.13) | 2.41 (1.81, 3.13) | 2.40 (1.82, 3.14) |

**Footnotes:** Variables are presented as median (interquartile range, IQR) or *n* (%).
Abbreviations: CVD, cardiovascular disease; HbA1c, glycosylated hemoglobin; HDL-C, HDL cholesterol; LDL-C, LDL cholesterol; METS-VF, Metabolic Score for Visceral Fat; CVAI, Chinese Visceral Adiposity Index; EVA, visceral adipose tissue area estimate; DAAT: deep abdominal adipose tissue; DAI: dysfunctional adiposity index; VAT: visceral adiposity index; LAP: lipid accumulation product.

### Association of VAT surrogates with outcomes

During a median follow-up of 20.13 years (IQR: 19.3–21.4), there were 4,068 deaths (4% of the total sample) attributable to CVD. Of these deaths, 2,938 (72.2%) were due to cardiac causes, 875 (21.5%) were stroke-related, and 255 (6.3%) were attributed to other cardiovascular causes. The cause-specific deaths for each sex are presented in **Supplementary Table 4** and **Supplementary Table 5.** For the primary outcome, each 1-SD increase in METS-VF (HR: 2.74 [95% CI 2.64–2.87]), EVA (HR: 2.20 [2.13–2.27]), CVAI (HR: 2.16 [2.09–2.23]), DAAT (HR: 1.50 [1.46–1.55]), and LAP (HR: 1.16 [1.13–1.20]) indices were positively associated with a higher hazard of CVD mortality. Conversely, DAI and VAI were not significantly associated with the primary outcome (**Figure 2A**). After adjusting for potential confounders, METS-VF (aHR: 1.17 [1.12–1.23]), EVA (aHR: 1.14 [1.10–1.19]), CVAI (aHR: 1.13 [1.09–1.18]), and DAAT (aHR: 1.13 [1.09–1.18]) remained associated with an increased hazard of the primary outcome, whereas LAP, DAI, and VAI showed no significant association (**Figure 2B**). Stratifying these analyses by cause-specific CVD causes yielded a similar trend, whereby the unadjusted models for METS-VF, EVA, CVAI, DAAT, and LAP were positively associated with cardiac, stroke, and other CVD-related deaths. However, in the adjusted models, only METS-VF predicted cardiac- and stroke-related deaths, whereas EVA, CVAI, and DAAT predicted stroke-related deaths (**Supplementary Table 6**).

**Figure 2:**
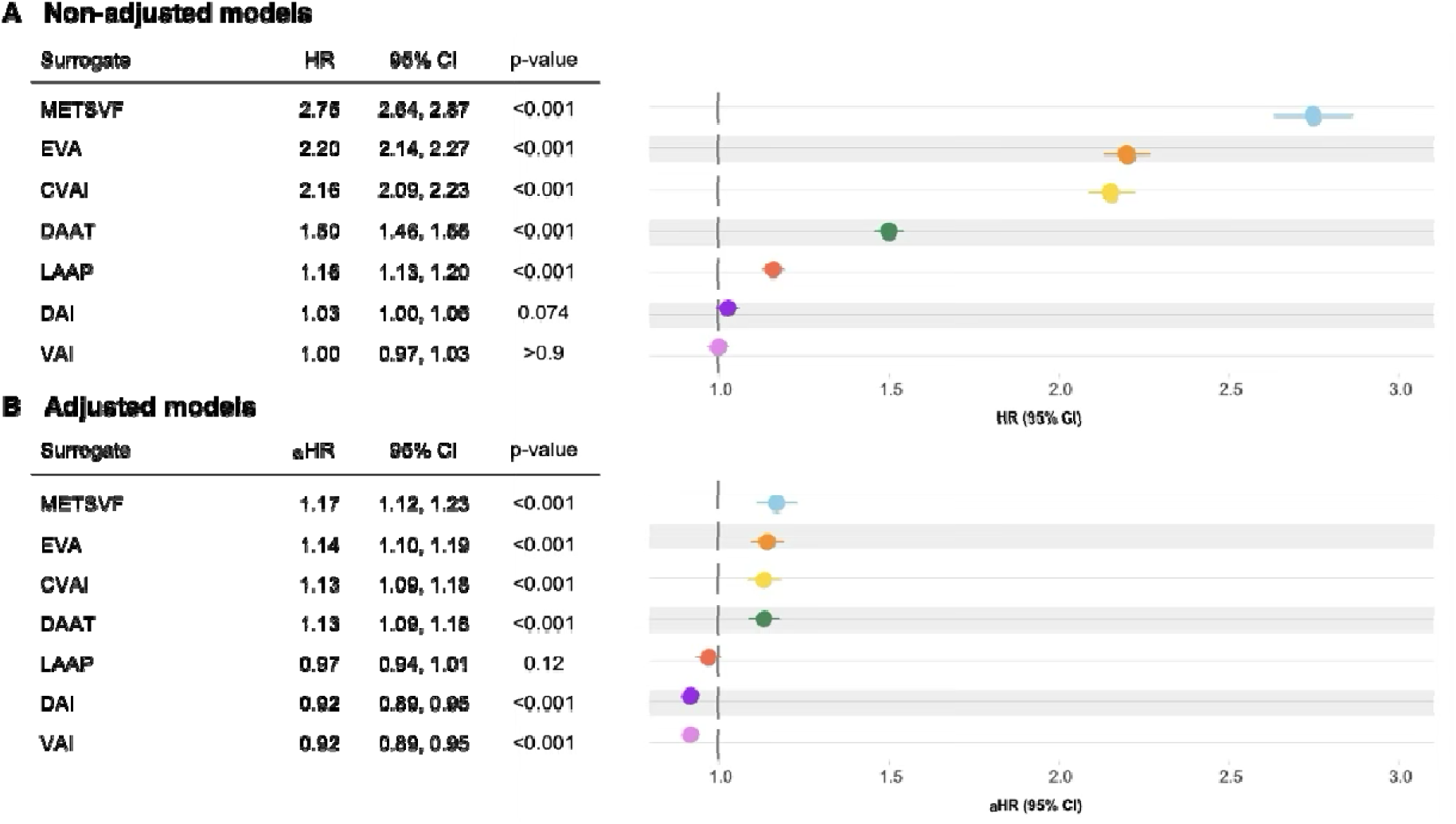
Comparison of the association of clinical surrogates estimating visceral fat for all CVD mortality causes using Cox proportional hazard regression models. **Footnotes:** Each circle represents the HR, and the line that crosses it is the 95% CI. **(A)** Association between clinical surrogates and all-cause cardiovascular mortality using unadjusted Cox regression models. **(B)** Association between clinical surrogates and all-cause cardiovascular mortality, using adjusted Cox regression models by age, sex, poor-quality diet, physical activity, and metabolic syndrome. Abbreviations: HR, hazard ratio; CI, confidence interval.

### Discrimination performance of VAT surrogates

The fixed-point discrimination for each surrogate revealed that METS-VF demonstrated the highest discrimination (AUROC: 0.722 [0.720–0.724]), followed by EVA (AUROC: 0.720 [0.718–0.722]), CVAI (AUROC: 0.702 [0.700–0.704]), DAAT (AUROC: 0.626 [0.624–0.629]), and LAAP (AUROC: 0.516 [0.514–0.518]). The DAI and VAI indices did not demonstrate significant discriminative ability for the overall outcome (**Figure 3A**). Time-dependent discrimination analyses displayed similar trends, whereas the METS-VF, EVA, and CVAI indexes maintained good discrimination over time, displaying AUROC values above 70% at most time points on the curve. DAAT showed moderate performance, with AUROC values between 60–65%, whereas LAAP, DAI, and VAI remained below 55%, indicating poor time-dependent discrimination. All indices tended to increase their discriminatory capacity after 15 years of follow-up (**Figure 3B**). Regarding the C-statistics, we found consistent results in which METS-VF and EVA both achieved a C-statistic of 0.72 [0.72–0.74], followed by CVAI (0.71 [0.71–0.72]) and DAAT (0.63 [0.62–0.64]).

**Figure 3:**
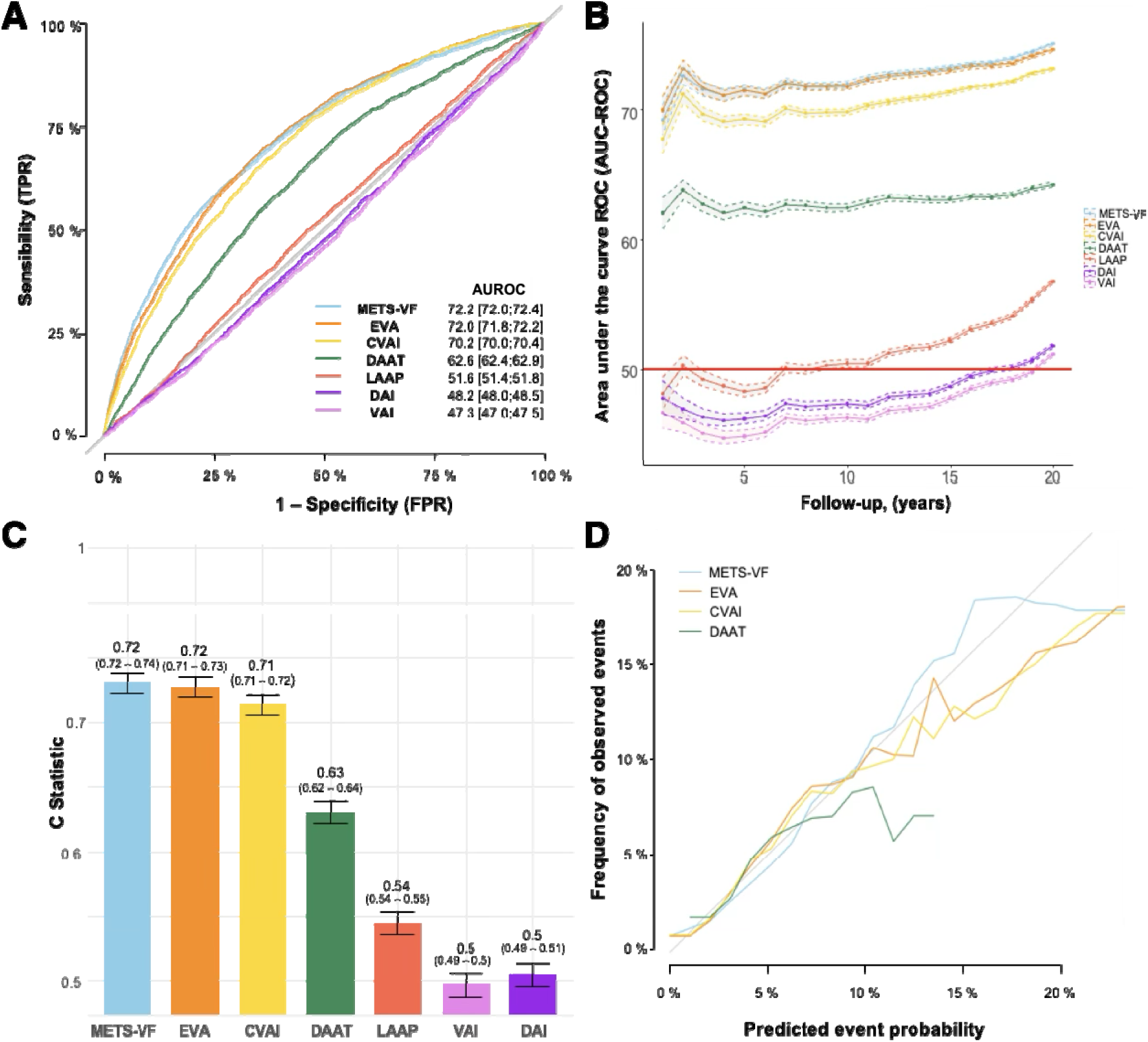
Predictive value of each surrogate for all CVD mortality causes. **Footnote:** **(A)** ROC curves showing the discriminative performance for mortality of each surrogate; AUC values and 95% confidence intervals are shown in percentage. **(B)** Time-dependent ROC curves illustrating the predictive performance of each surrogate over a 20-year follow-up period. Each circle represents one year of follow-up; shaded areas indicate 95% confidence intervals. **(C)** C-statistics with 95% confidence intervals for each surrogate. **(D)** Calibration curves for the best-performing surrogates across 20 years of follow-up. Abbreviations: TPR, true positive rate; FPR, false positive rate; AUC, area under the curve.

Conversely, LAAP, DAI, and VAI had C-statistics below 0.55 (**Figure 3C**). For model calibration, METS-VF demonstrated the best agreement between predicted and observed CVD mortality events. EVA and CVAI showed good calibration but tended to overpredict mortality in the lower-risk range, whereas DAAT exhibited mild overprediction at higher estimated risk values (**Figure 3D**). A similar trend was observed for cardiac-related mortality (**Supplementary Figure 2**), stroke-related mortality (**Supplementary Figure 3**), and mortality due to other CVD causes (**Supplementary Figure 4**), although for the latter there were insufficient events to evaluate calibration.

### Thresholds of VAT surrogates to predict CVD and cause-specific mortality

We derived and applied optimal cut-off points for the seven VAT surrogates and assessed their discrimination performance for predicting CVD mortality. In the adjusted Cox models, four surrogates showed a positive association with increased risk of this outcome. Participants with a METS-VF value >7.33 (aHR: 1.20 [1.12–1.29]) or a CVAI score >137.97 (aHR: 1.20 [1.11–1.29]) had, on average, a 20% higher hazard of CVD mortality after accounting for the effect of confounders compared with those at or below these thresholds. Furthermore, individuals with an EVA score >138.94 had a 24% higher hazard (aHR: 1.24 [1.16–1.34]), and a DAAT value >2.64 conferred a 15% higher hazard to experience the primary outcome (aHR, 1.15 [1.07–1.23]) compared with participants below each threshold (**Table 2**).

**Table 2:** Thresholds for each surrogate included in the analysis and their association with CVD mortality.

| Surrogate | Threshold | HR (95% IC) | aHR (95% IC) | C Statistic (95% CI) |
| --- | --- | --- | --- | --- |
| METS-VF | 7.33 | 4.4 (4.14 – 4.68) | 1.2 (1.12 – 1.29) | 0.66 (0.65-0.66) |
| EVA | 138.94 | 4 (3.8 – 4.3) | 1.24 (1.16 – 1.34) | 0.66 (0.66-0.67) |
| CVAI | 137.97 | 3.71 (3.48 – 3.96) | 1.2 (1.11 – 1.29) | 0.66 (0.65-0.67) |
| DAAT | 153.41 | 2.4 (2.24 – 2.56) | 1.32 (1.22-1.42) | 0.6 (0.6-0.61) |
| LAAP | 44.66 | 2.4 (2.24 - 2.56) | 1.32 (1.22 – 1.42) | 0.54 (0.53-0.54) |
| VAI | 2.22 | 1.37 (1.28 – 1.45) | 0.98 (0.91 – 1.1) | 0.51 (0.5-0.51) |
| DAI | 1.03 | 1.03 (0.95 – 1.11) | 0.94 (0.86 – 1.02) | 0.51 (0.5-0.51) |

**Footnotes:** Variables' cut-off points are presented in their units.
The optimal cut-off point was determined by maximization of the log-rank statistic. C-statistic and 95% CI for each surrogate were calculated using Cox regression models.
Abbreviations: CI, confidence interval

### Added performance of the METS-VF index to Globorisk prediction

As a complementary analysis, we compared the performance of the Globorisk model with and without the addition of METS-VF, given that it was the index with the highest discrimination overall and across risk strata (**Supplementary Table 7**). Adding METS-VF improved overall discrimination (ΔC-statistic = 0.003) and discrimination within risk strata; notably, gains were larger in the intermediate-risk (ΔC-statistic = 0.042) and very-low-risk groups (ΔC-statistic = 0.016), which may be relevant for early identification of high-risk individuals who might otherwise be misclassified.

## DISCUSSION

In this study of Mexican adults enrolled in the MCPS and followed for a median of 20 years, we evaluated the performance of seven VAT-related clinical surrogates in predicting CVD and CVD cause-specific mortality. We found that METS-VF, EVA, CVAI, and DAAT were positively associated with a higher hazard of the evaluated outcomes. Consistently, these four surrogates remained as the best VAT-surrogates to predict these outcomes, with good discrimination and calibration capacity, for which we derived specific thresholds for each outcome. Finally, we found that, when adding METS-VF to GloboRisk, discrimination improved for individuals in the intermediate-risk group. Beyond confirming previously reported individual associations of each VAT-surrogate, this study extends existing evidence by evaluating multiple VAT clinical surrogates simultaneously in a large prospective cohort with long-term follow-up and CVD outcomes.

Our findings highlight the potential utility of VAT-related clinical surrogates as clinical tools to estimate VAT among patients at increased risk of CVD mortality. Given that the effect of VAT on cardiovascular health represents an “invisible” burden, given that this compartment is not adequately captured by traditional anthropometric measures, VAT estimations are increasingly recognized as a valuable component to assess an individual’s cardiometabolic risk. Hence, VAT-related clinical surrogates offer an accessible, simple, low-cost, and scalable approach that can be incorporated into primary care settings to detect individuals who might have high VAT despite having been classified as overweight or normal weight based solely on BMI or WC, particularly in LMICs where direct image-based methods are not fully available ^44^. These strategies could be combined with individual-level efforts aimed at reducing excess adiposity and improving metabolic health, including structured aerobic-plus-resistance exercise programs, dietary patterns emphasizing whole grains, low sodium, and high fiber intake, and selected pharmacotherapies with demonstrated effects on total fat reduction (e.g., GLP-1 receptor agonists, SGLT2 inhibitors) ^45^. Beyond individual-level interventions, public-health interventions that promote healthy food environments, active mobility, and equitable access to preventive care are essential to achieve sustained reductions in VAT and related cardiometabolic diseases. Furthermore, given the substantial heterogeneity in patients who may benefit from VAT assessment, there is a need to define and prospectively validate national-representative and clinically meaningful thresholds for high VAT accumulation, ideally using percentile-based references stratified by sex, age, and ethnicity, to facilitate the identification of abnormal VAT burden and support more precise risk assessment across diverse clinical settings. Overall, these strategies may help improve cardiometabolic health in at-risk populations in LMICs.

Regarding METS-VF, we found that this index has the strongest predictive capacity for CVD mortality. These findings align with previous evidence from prospective cohorts, where higher METS-VF values were consistently associated with a higher risk of CVD mortality. For example, a study by Zhu *et al.* in adults over 40 years from the Cardiometabolic and Cancer Cohort Study in China demonstrated that participants in the highest quartile of METS-VF had substantially higher hazards of incident CVD and all-cause mortality compared with individuals in lowest quartile. Furthermore, in the same study, METS-VF also demonstrated superior discrimination capacity in all participants (AUC: 0.611, 0.601 – 0.620), normal glucose tolerance (AUC: 0.642, 0.625–0.657 groups compared with other evaluated obesity and insulin resistance clinical surrogates, such as METS-IR, HOMA-IR, VAI, BMI and WtHR ^46^. Similarly, Tan *et al.* found that, among middle-aged and older adults from the China Health and Retirement Longitudinal Study cohort, METS-VF was positively associated with a higher risk of developing incident stroke and a composite CVD event, and that triglycerides also partially mediated this association ^47^. Finally, in a study by Liu *et al.* in adults from the Kailuan Study cohort, they found that a cumulative exposure to elevated METS-VF over six years predicted the incidence of a composite of CVD events and all-cause mortality. In the same study, cumulative exposure to METS-VF outperformed cumulative BMI, waist circumference, and WtHR in discrimination capacity, being a more reliable surrogate than these traditional measures to predict CVD ^48^. The magnitude and direction of the associations observed in our cohort are consistent with those reported in these prospective studies, suggesting similar predictive performance across populations. Importantly, the thresholds identified in this study were derived specifically from a Mexican adult cohort, highlighting the relevance of population-specific calibration when applying VAT-related surrogates in clinical practice.

Prior studies using clinical VAT surrogates have explored the association between these indices (CVAI, EVA, DAAT, LAP and DAI) and incident CVD events in different populations. For example, Ren *et al.* and Kouli *et al.* demonstrated in prospective cohorts that higher values of CVAI were positively associated with a higher risk of incident CVD, heart disease, and stroke among Chinese populations over 45 years of age ^49,50^. Similarly, Kahn *et al.* and Amato *et al.* showed that the LAP and VAI indices were positively associated with adverse cardiometabolic profiles, including metabolic syndrome, diabetes, high blood pressure, high LDL-C and total cholesterol levels, low HDL-C levels, and high triglycerides levels. Furthermore, these indexes independently demonstrated a good predictive capacity for CVD or all-cause mortality in different population-based cohorts ^35,36,51^. For EVA and DAAT, the evidence is limited, as these indices have not been fully explored as predictors of incident CVD events or mortality. However, Golabi *et al.* and Garofallo *et al.* demonstrated that higher values of EVA and DAAT correlated with an adverse cardiometabolic profile, such as higher levels of LDL-C, HDL-C, and serum triglycerides ^34,52,53^. Our findings complement and expand these studies, as they provide a large evaluation in a Mexican adult cohort of multiple VAT surrogates, especially EVA, CVAI, and DAAT, for which the association with and prediction of CVD mortality had not been previously evaluated. In contrast to prior studies that mainly focused on cardiometabolic profiles or incident events, our study provides longitudinal evidence linking these surrogates to CVD mortality. Future research should evealuate wheter longitudinal changes in clinical VAT indices add prognostic value beyond baseline measurements and outperform traditional anthropometric indices, including BMI and waist circumference. Moreover, their integration into multivariable risk prediction models should be assessed to determine improvements in discrimination, calibration, and risk reclassification, thereby informing their potential role in clinical risk stratification, preventive guidelines, and the early identification of patients at risk for CVD across different visceral adiposity phenotypes, including normal-weight visceral obesity.

### Strengths and limitations

Several strengths and limitations should be considered when interpreting our findings. A major strength of this study is the large sample of middle-aged Mexican adults with a median follow-up of approximately 20 years, which allowed the assessment of long-term CVD mortality using complete anthropometric and biochemical characterization. However, several limitations should be considered. First, the association and predictive capacity of these surrogates could not be evaluated in individuals younger than 35 years, due to restrictions in the original MCPS cohort inclusion criteria ^27^. This is relevant, given that previous evidence has shown that VAT accumulation at younger ages could contribute to longer exposure to VAT-related adverse outcomes ^54^. Second, although we compared the added performance of METS-VF versus Globorisk to predict CVD mortality, previous studies in MCPS have demonstrated that the predictive capacity of CVD risk scores is modest, even after recalibration in this cohort ^55^. Hence, the added performance of VAT should be interpreted as a modest increment in the predictive capacity for this outcome. Third, our analyses were restricted only to CVD mortality, leading to uncertainty regarding which VAT-related clinical surrogate could better predict non-fatal CVD outcomes. Fourth, MCPS is restricted to only two urban districts of Mexico City (i.e., Coyoacán and Iztapalapa), which may not be fully representative of broader national populations or rural settings where sociodemographic and clinical determinants may shift the distribution of VAT. Additionally, the marked sex imbalance in the cohort (approximately 67% women and 33% men) may have limited the precision of sex-specific estimates, particularly in men, leading to an underestimation of the predictive performance of VAT surrogates in this group. Furthermore, the lack of information on menopausal status precluded stratified analyses in women, which is relevant given the well-established impact of menopause on visceral fat accumulation and cardiometabolic risk. Finally, although our study found that four specific VAT-related surrogates showed the highest predictive capacity for CVD mortality, they are not intended to replace imaging-based methods such as CT, MRI, or DXA, which remain the gold standard for VAT quantification. This is relevant, as these indices are not well calibrated for specific populations, such as pediatric and very elderly groups, which leads to an area of opportunity to develop indices specifically tailored for these populations.

## CONCLUSIONS

In this sub analysis of a large prospective study including ∼100,000 Mexican adults followed for long-term CVD mortality, we found that four previously developed VAT-related clinical surrogates (METS-VF, EVA, CVAI, and DAAT) were positively associated with, and showed the highest predictive capacity for, all-cause and CVD-specific mortality. Specifically, adding METS-VF to a cardiovascular risk score among individuals classified as intermediate-risk improved discrimination for these outcomes. Our findings support the view that the use of any of these four surrogates may be a useful strategy to enable early identification of individuals with high VAT accumulation and, consequently, high cardiometabolic risk.

## Data Availability

Data from the Mexico City Prospective Study are available to bona fide researchers. The study Data and Sample Sharing policy can be viewed (in English or Spanish) on the MCPS website (https://www.ctsu.ox.ac.uk/research/mcps). Available study data can be examined in detail through the study Data Showcase (https://datashare.ndph.ox.ac.uk/mexico). The code and Supplementary Material are publicly available in a GitHub repository and can be accessed at: https://github.com/neftalivilla/VAT_INDEXES_MCPS.

## ABBREVIATIONS

CVD: Cardiovascular disease
LMICs: Low and middle-income countries
VAT: Visceral adipose tissue
BMI: Body mass index
WC: Waist circumference
MCPS: Mexico City Prospective Study
METS-VF: Metabolic score for visceral fat
EVA: Estimate of visceral adipose tissue area
CVAI: Chinese visceral adiposity index
DAAT: Deep abdominal adipose tissue
LAP: Lipid accumulation product
VAI: Visceral adiposity index
DAI: Dysfunctional adiposity index
SD: Standard deviation
WtHR: Waist-to-height ratio
HRs: Hazard ratios
AUROC: Area under the receiver operating characteristic curve
AUC: Area under the curve

## Ethics approval and consent to participate

The study was approved by Ethics Committees at the Mexican Ministry of Health, the Mexican National Council for Science and Technology, and the University of Oxford, UK. All participants provided written informed consent.

## Consent for publication

Not applicable.

## Availability of data and materials

Data from the Mexico City Prospective Study are available to bona fide researchers. The study’s Data and Sample Sharing policy can be viewed (in English or Spanish) on the MCPS website (https://www.ctsu.ox.ac.uk/research/mcps). Available study data can be examined in detail through the study’s Data Showcase (https://datashare.ndph.ox.ac.uk/mexico). The code and Supplementary Material is publicly available in a GitHub repository and can be accessed at: https://github.com/neftalivilla/VAT_INDEXES_MCPS.

## Authors’ contributions

Research idea and study design: JEML, OYBC, NEAV

Data acquisition: JBC, PKM, RTC, JAD

Data analysis/interpretation: JEML, OYBC, NEAV

Statistical analysis: JEML, OYBC, NEAV

Manuscript drafting: JEML, MFSV, MCTC, GCCS, JRB, JS, CAFM, OYBC, NEAV

Supervision or mentorship: OYBC, NEAV

Each author contributed important intellectual content during manuscript drafting or revision and accepted accountability for the overall work by ensuring that questions about the accuracy or integrity of any portion of the work were appropriately investigated and resolved.

## Acknowledgements

This research has been conducted using Mexico City Prospective Study (MCPS) Data under Application Number 2022-012-08. The MCPS (https://www.ctsu.ox.ac.uk/research/mcps) is a long-standing scientific collaboration between researchers at the National Autonomous University of Mexico and the University of Oxford. Carlos A. Fermín-Martínez is in the PECEM Program of the Faculty of Medicine at UNAM. CAFM is supported by CONACyT.

