## Supplementary Material for "Predictive performance of seven clinical surrogates of visceral adipose tissue for cardiovascular mortality: A sub-analysis of 102,385 adults from the Mexico City Prospective Study"

**Index**

### Supplementary methods

Supplementary Figure 1: Direct acyclic graph to evaluate potential mediators, confounders, interactors, and instrumental variables related to the relation between clinical surrogates that estimate visceral adipose tissue and cardiovascular disease mortality.

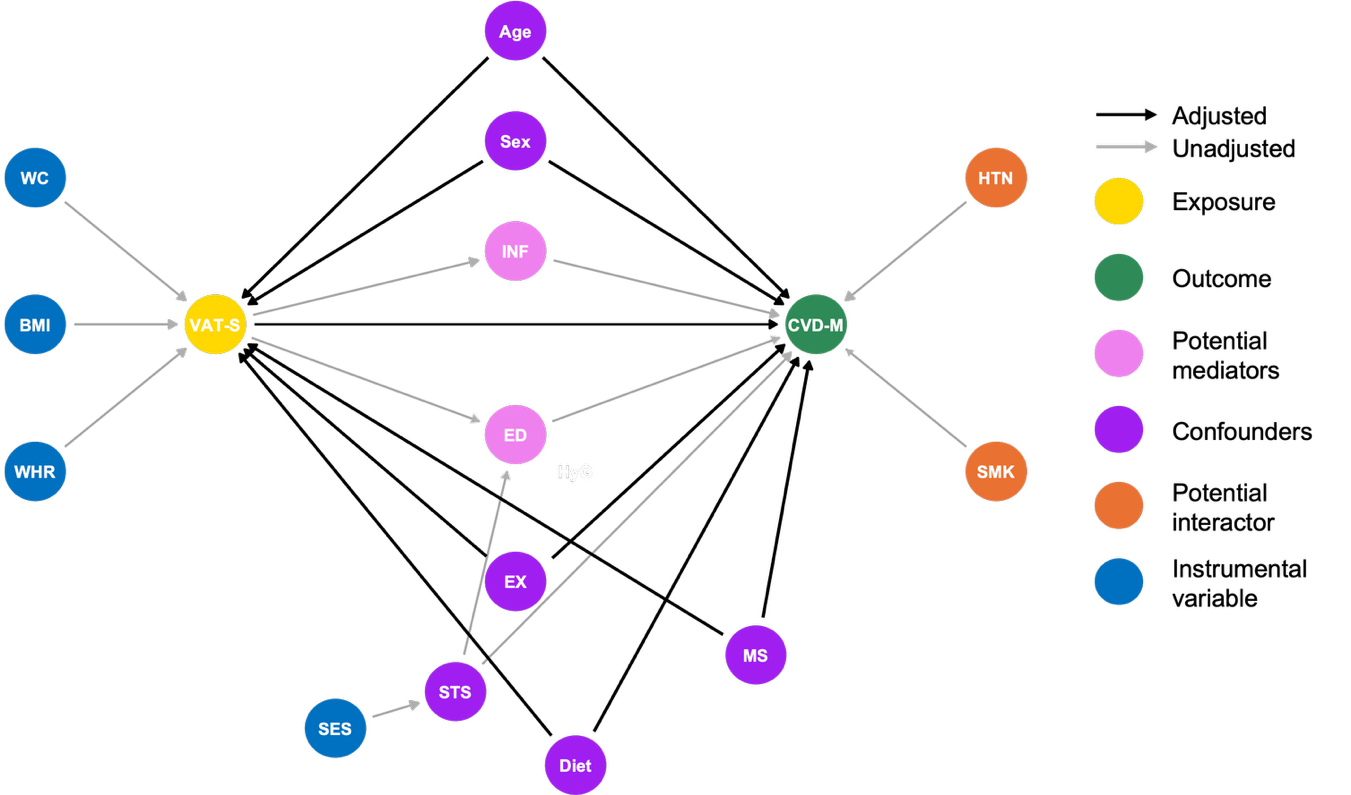

Abbreviations: WC, waist circumference; BMI, body mass index; WHR, waist-to-height ratio; VAT-S, surrogates that estimate visceral adipose tissue; SES, socio-economic status; INF, inflammation; ED, endothelial dysfunction; EX, exercise; MS, metabolic syndrome; CVD-M, cardiovascular disease mortality; HTN, Hypertension; SMK, smoking.

Supplementary Figure 2. Predictive value of each surrogate for cardiac-related mortality

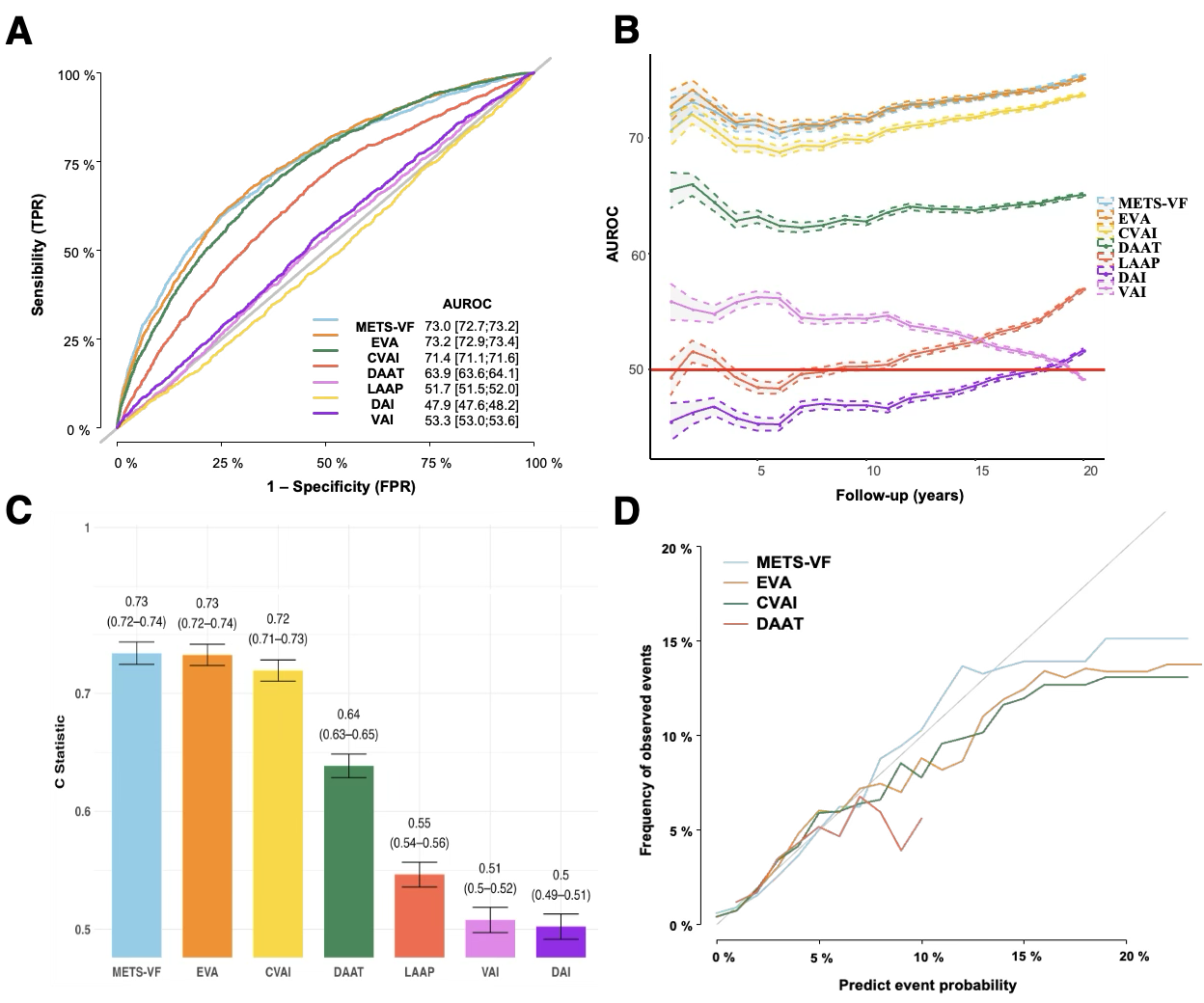

Footnotes:

(A) ROC curves showing the discriminative performance for mortality of each surrogate; AUC values and 95% confidence intervals are shown in percentage.
(B) Time-dependent ROC curves illustrating the predictive performance of each surrogate over a 20-year follow-up period. Each circle represents one year of follow-up; shaded areas indicate 95% confidence intervals.
(C) C-statistics with 95% confidence intervals for each surrogate.
(D) Calibration curves for the best-performing surrogates across 20 years of follow-up.

Abbreviations: TPR, true positive rate; FPR, false positive rate; AUC, area under the curve.

Supplementary Figure 3. Predictive value of each surrogate for stroke-related mortality

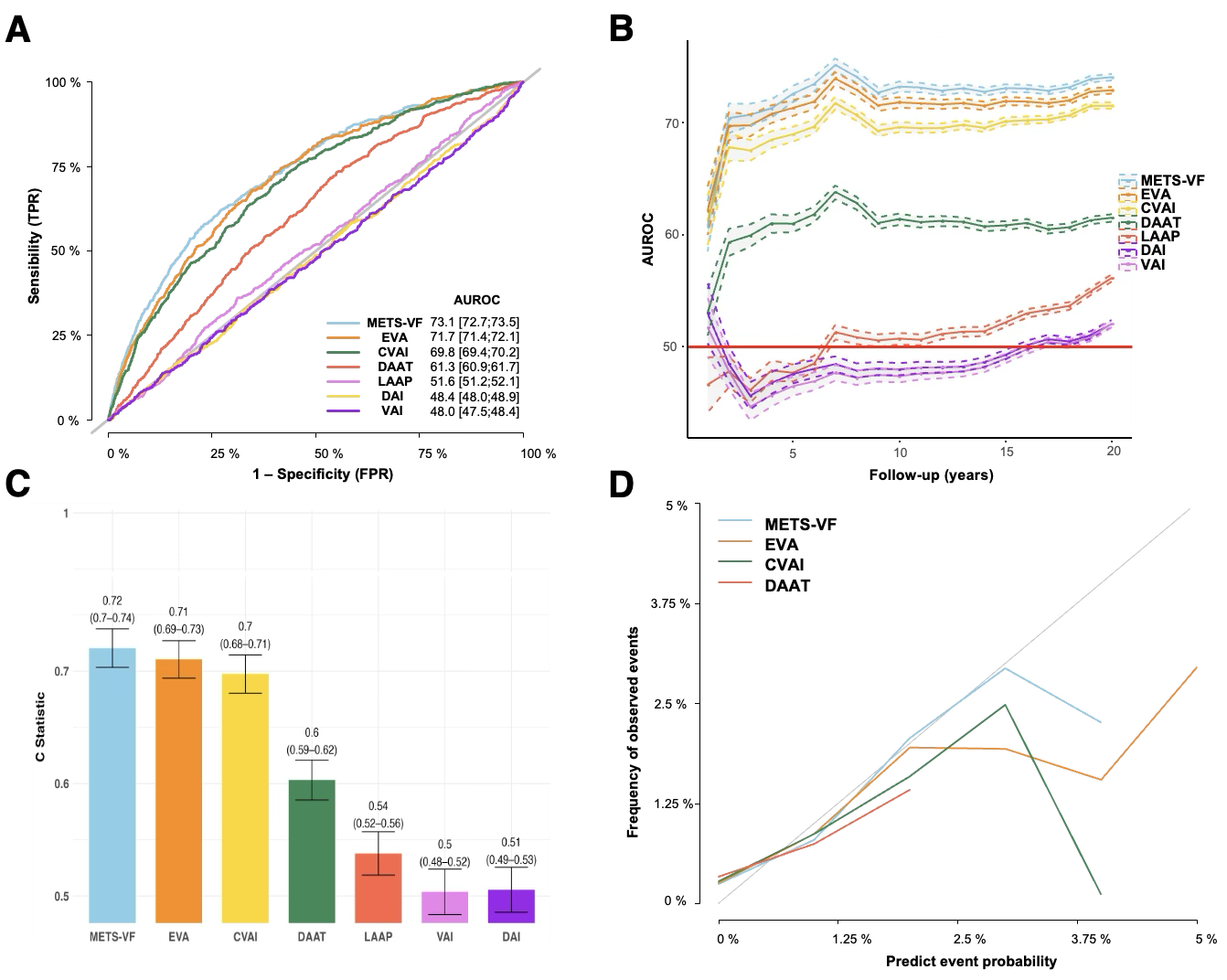

Footnotes:

(A) ROC curves showing the discriminative performance for mortality of each surrogate; AUC values and 95% confidence intervals are shown in percentage.
(B) Time-dependent ROC curves illustrating the predictive performance of each surrogate over a 20-year follow-up period. Each circle represents one year of follow-up; shaded areas indicate 95% confidence intervals.
(C) C-statistics with 95% confidence intervals for each surrogate.
(D) Calibration curves for the best-performing surrogates across 20 years of follow-up.

Abbreviations: TPR, true positive rate; FPR, false positive rate; AUC, area under the curve.

Supplementary Figure 4. Predictive value of each surrogate for other causes of mortality

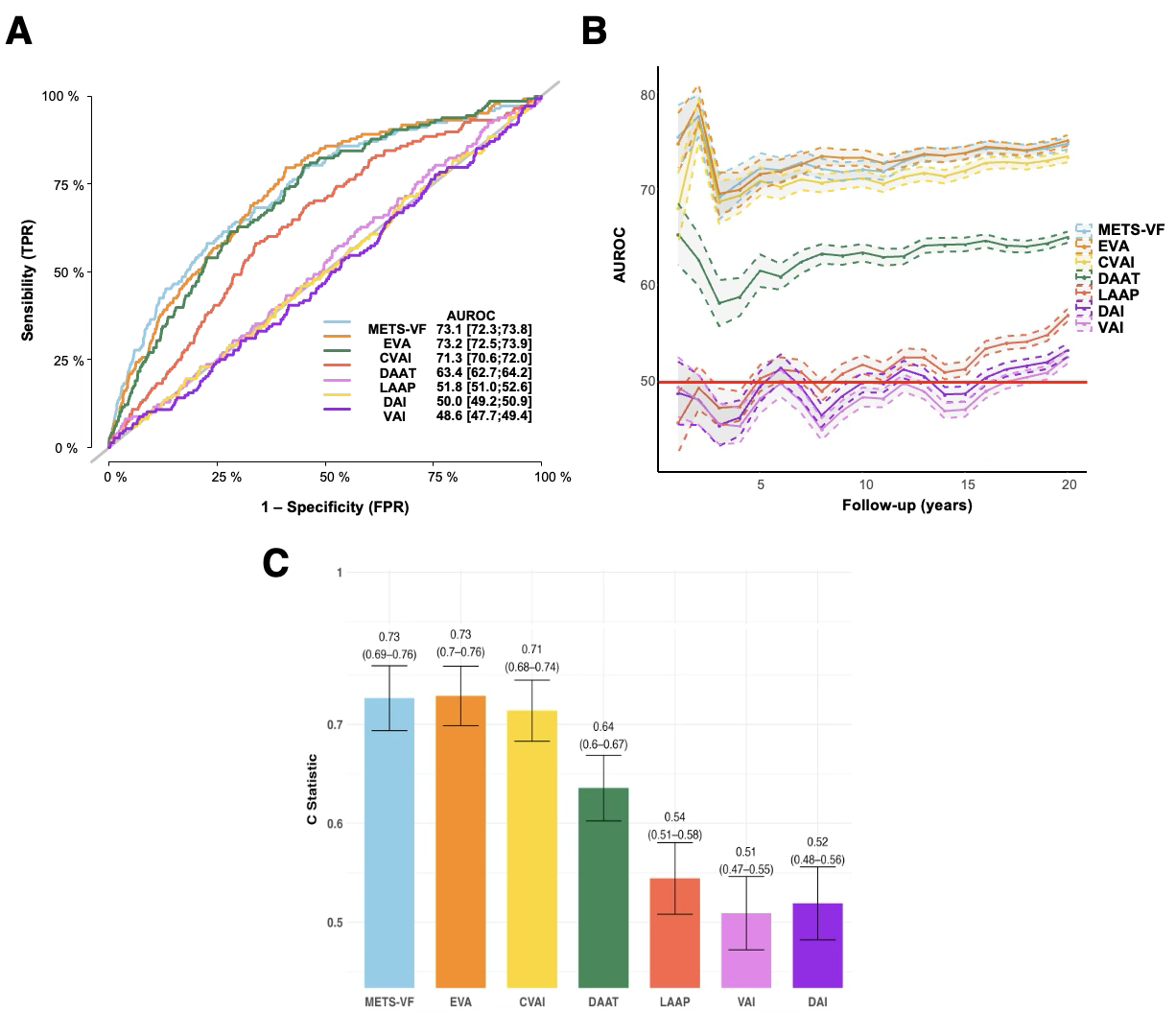

Footnotes: The calibration curve could not be performed due to insufficient variability in predicted risks, likely caused by a small number of events for this outcome.

(A) ROC curves showing the discriminative performance for mortality of each surrogate; AUC values and 95% confidence intervals are shown in percentage.
(B) Time-dependent ROC curves illustrating the predictive performance of each surrogate over a 20-year follow-up period. Each circle represents one year of follow-up; shaded areas indicate 95% confidence intervals.
(C) C-statistics with 95% confidence intervals for each surrogate.

Abbreviations: TPR, true positive rate; FPR, false positive rate; AUC, area under the curve.

Supplementary Table 1: STROBE cohort guidelines report for the study. This checklist was completed on [xxx] using GoodReports (developed by the EQUATOR Network in collaboration with Penelope.ai). The STROBE statement was followed for reporting observational studies (von Elm E, Altman DG, Egger M, et al. The Strengthening the Reporting of Observational Studies in Epidemiology (STROBE) Statement: guidelines for reporting observational studies).

|  | Item No | Recommendation | Page Number |
| --- | --- | --- | --- |
| **Title and abstract** | 1 | (*a*) Indicate the study’s design with a commonly used term in the title or the abstract |  |
|  |  | (*b*) Provide in the abstract an informative and balanced summary of what was done and what was found |  |
| Introduction | | |  |
| Background/rationale | 2 | Explain the scientific background and rationale for the investigation being reported |  |
| Objectives | 3 | State specific objectives, including any prespecified hypotheses |  |
| Methods | | |  |
| Study design | 4 | Present key elements of study design early in the paper |  |
| Setting | 5 | Describe the setting, locations, and relevant dates, including periods of recruitment, exposure, follow-up, and data collection |  |
| Participants | 6 | (*a*) Give the eligibility criteria, and the sources and methods of selection of participants. Describe methods of follow-up |  |
|  |  | (*b*) For matched studies, give matching criteria and number of exposed and unexposed |  |
| Variables | 7 | Clearly define all outcomes, exposures, predictors, potential confounders, and effect modifiers. Give diagnostic criteria, if applicable |  |
| Data sources/ measurement | 8* | For each variable of interest, give sources of data and details of methods of assessment (measurement). Describe comparability of assessment methods if there is more than one group |  |
| Bias | 9 | Describe any efforts to address potential sources of bias |  |
| Study size | 10 | Explain how the study size was arrived at |  |
| Quantitative variables | 11 | Explain how quantitative variables were handled in the analyses. If applicable, describe which groupings were chosen and why |  |
| Statistical methods | 12 | (*a*) Describe all statistical methods, including those used to control for confounding |  |
|  |  | (*b*) Describe any methods used to examine subgroups and interactions |  |
|  |  | (*c*) Explain how missing data were addressed |  |
|  |  | (*d*) If applicable, explain how loss to follow-up was addressed |  |
|  |  | (*e*) Describe any sensitivity analyses |  |
| Results | | |  |
| Participants | 13* | (a) Report numbers of individuals at each stage of study—eg numbers potentially eligible, examined for eligibility, confirmed eligible, included in the study, completing follow-up, and analysed |  |
|  |  | (b) Give reasons for non-participation at each stage |  |
|  |  | (c) Consider use of a flow diagram |  |
| Descriptive data | 14* | (a) Give characteristics of study participants (eg demographic, clinical, social) and information on exposures and potential confounders |  |
|  |  | (b) Indicate number of participants with missing data for each variable of interest |  |
|  |  | (c) Summarise follow-up time (eg, average and total amount) |  |
| Outcome data | 15* | Report numbers of outcome events or summary measures over time |  |
| Main results | 16 | (*a*) Give unadjusted estimates and, if applicable, confounder-adjusted estimates and their precision (eg, 95% confidence interval). Make clear which confounders were adjusted for and why they were included |  |
|  |  | (*b*) Report category boundaries when continuous variables were categorized |  |
|  |  | (*c*) If relevant, consider translating estimates of relative risk into absolute risk for a meaningful time period |  |
| Other analyses | 17 | Report other analyses done—eg analyses of subgroups and interactions, and sensitivity analyses |  |
| Discussion | | |  |
| Key results | 18 | Summarise key results with reference to study objectives |  |
| Limitations | 19 | Discuss limitations of the study, taking into account sources of potential bias or imprecision. Discuss both direction and magnitude of any potential bias |  |
| Interpretation | 20 | Give a cautious overall interpretation of results considering objectives, limitations, multiplicity of analyses, results from similar studies, and other relevant evidence |  |
| Generalisability | 21 | Discuss the generalisability (external validity) of the study results |  |
| Other information | | |  |
| Funding | 22 | Give the source of funding and the role of the funders for the present study and, if applicable, for the original study on which the present article is based |  |

Supplementary Table 2. ICD-10 codes for cardiovascular disease mortality and the number of deaths for all specific codes.

| Cause specific-mortality and number of deaths | ICD-10 codes (n) |
| --- | --- |
| Cardiac deaths  n = 2,938 | *I050 (3), I051 (1), I059 (13), I060 (1), I070 (1), I071 (3), I079 (1), I080 (5), I081 (1), I091 (1), I099 (14), I110 (125), I119 (27), I200 (2), I209 (5), I210 (4), I211 (8), I213 (2), I219 (2128), I220 (1), I221 (1), I229 (1), I248 (1), I249 (58), I251 (49), I255 (3), I258 (7), I259 (114), I270 (7), I272 (4), I279 (6), I301 (1), I318 (1), I330 (4), I340 (5), I350 (11), I351 (1), I358 (3), I38X (10), I420 (13), I426 (1), I429 (2), I441 (1), I442 (2), I443 (3), I459 (1), I469 (3), I471 (3), I472 (2), I482 (1), I489 (6), I48X (11), I490 (5), I499 (19), I500 (50), I501 (9), I509 (107), I518 (4), I519 (8), Q210 (1), Q231 (1), Q238 (1), R570 (51)* |
| Cerebrovascular deaths  n = 875 | *F019 (10), G459 (1), I600 (2), I602 (1), I608 (1), I609 (78), I610 (1), I613 (1), I614 (1), I615 (2), I618 (1), I619 (211), I620 (10), I629 (8), I633 (3), I634 (25), I635 (8), I638 (1), I639 (85), I64X (111), I671 (8), I672 (1), I674 (6), I678 (117), I679 (121), I691 (1), I693 (11), I694 (15), I698 (34)* |
| Other cardiovascular deaths  n = 255 | *E115 (4), E145 (5), I260 (1), I269 (76), I709 (2), I710 (3), I712 (1), I713 (5), I714 (7), I718 (1), I719 (2), I720 (1), I729 (2), I739 (5), I741 (1), I771 (9), I779 (1), I802 (8), I803 (1), I822 (1), I828 (2), I829 (2), I839 (1), I872 (6), I879 (1), I888 (1), I99X (3), K550 (88), K559 (14), K761 (1)* |

Footnote:

Abbreviations: ICD 10, International Statistical Classification of Diseases and Related Health Problems, Tenth Revision.

| **Autor (year)** | **Surrogate name** | **Formulae** |
| --- | --- | --- |
| Brudavani,V. et al. (2006) | Deep abdominal adipose tissue (DAAT) | Men  $-382.9+\left( 1.09 \times weight \left[ kg \right] \right)+\left( 6.04 \times WC \left[ cm \right] \right)-\left( 2.29 \times BMI[\frac{kg}{m^{2}}] \right)$  Women  $-278-\left( 0.86 \times weight \left[ kg \right] \right)+\left( 5.19 \times WC \left[ cm \right] \right)$ |
| Kahn, H.S., & Valdez, R. (2003) | The lipid accumulation product (LAP) | Men  $\left( WC \left[ cm \right]-65 \right)\times\left( Tg \left[ \frac{mmol}{L} \right] \right)$  Women  $\left( WC \left[ cm \right]-58 \right)\times\left( Tg \left[ \frac{mmol}{L} \right] \right)$ |
| Amato, M.C. et al. (2010) | Visceral adiposity index (VAI) | Men  $\frac{WC\left[ cm \right]}{\left( 3.98+\left( 1.88xBMI[\frac{kg}{m^{2}}] \right) \right)} \times\frac{Tg \left[ \frac{mmol}{L} \right]}{1.03} \times\frac{1.31}{HDLC\left[ \frac{mmol}{L} \right]}$  Women  $\frac{WC \left[ cm \right]}{\left( 36.85+\left( 1.89\times BMI[\frac{kg}{m^{2}}] \right) \right)} \times\frac{Tg \left[ \frac{mmol}{L} \right]}{0.81} \times\frac{1.52}{HDLC \left[ \frac{mmol}{L} \right]}$ |
| Wander, P.L. (2018) | Estimate of visceral adipose tissue area (EVA) | Men  $\left( 1.28 \times\mathrm{Age} \left[ \mathrm{years} \right] \right)+\left( 4.12 \times WC \left[ cm \right] \right)-\left( 0.53 \times HDLC \left[ \frac{mg}{dL} \right] \right)+\left( 0.14 \times Glucose \left[ \frac{mg}{dL} \right] \right)-319.5$  Women  $\left( 1.26\times Age\left[ years \right] \right)+\left( 1.89\times BMI[\frac{kg}{m^{2}}] \right)+\left( 2.16\times WC\left[ cm \right] \right)-$  $(HDLC[\frac{mg}{dL}])+(0.11\times LDLC[\frac{mg}{dL}])+(0.18\times Glucose[\frac{mg}{dL}])-207.22$ |
| Bello-Chavolla, O.Y. (2019) | Metabolic score for visceral fat (METS-VF) | METS-VF  $4.466+0.11+\left( \log\left( METS.IR \right) \right)^{3}+3.299\times\left( \log\left( WHR \right) \right)^{3}$  $+0.319\times\left( Male \right)+0.594\times\left( \log\left( Age\left[ years \right] \right) \right)$  Where METS.IR  $log(\left( 2\times Glucose\left[ \frac{mg}{dL} \right] \right)+(Tg\left[ \frac{mg}{dL} \right])\times$  $BMI[\frac{kg}{{mt}^{2}}]/(\log\left( HDLC\left[ \frac{mg}{dL} \right] \right))$ |
| Reyes-Barrera, J. (2021) | Dysfunctional adiposity index (DAI) | Men  $\left( \frac{WC [cm]}{22.79+2.68\times BMI[\frac{kg}{m^{2}}]} \right) \times\left( \frac{Tg[\frac{mmol}{L}]}{1.37} \right) \times\left( \frac{1.19}{HDLC[\frac{mmol}{L}]} \right)$  Women  $\left( \frac{WC [cm[}{24.02+2.37\times BMI[\frac{kg}{m^{2}}]} \right)\times\left( \frac{Tg[\frac{mmol}{L}]}{1.32} \right)\times\left( \frac{1.43}{HDLC[\frac{mmol}{L}]} \right)$ |
| Xia, MF., Chen, Y., Lin, HD. et al. (2016) | Chinese viseral adiposity index (CVAI) | Men  $-267.93+0.68\times Age\left[ years \right]+0.03\times BMI\left[ \frac{kg}{m^{2}} \right]+4.00\times$  $WC\left[ cm \right]+22.0\times(\log\left( Tg\left[ \frac{mmol}{L} \right] \right)-16.32\times HDLC[\frac{mmol}{L}]$  Women  $-187.32+1.71\times Age\left[ years \right]+4.23\times BMI\left[ \frac{kg}{m^{2}} \right]+1.12\times$  $WC\left[ cm \right]+39.76\times(log(Tg\left[ \frac{mmol}{L} \right]-11.66\times HDLC[\frac{mmol}{L}]$ |

Supplementary Table 3. Formulas, author, and year of publication for each surrogate that estimates visceral adipose tissue

Abbreviations: WC, waist circunference; BMI, body mass index; Tg, triglycerides; HDLC, cholesterol HDL; LDLC, cholesterol LDL; WHR, waist-to-height ratio.

Supplementary Table 4. Baseline characteristics of male patients included from the Mexico City Prospective Cohort, stratified according to the present outcome

| Characteristics | Overall male  n = 33,518 | Outcome | |
| --- | --- | --- | --- |
|  |  | **Censored** n = 31,761 | **Mortality due CVD** n = 1,751 |
| *Demographics* | | | |
| Age, (years) | 48 (40, 59) | 47 (40, 57) | 65 (54, 72) |
| Social development index, (%) |  |  |  |
| Very low | 8,999 (28%) | 8,578 (28%) | 421 (25%) |
| Low | 14,161 (44%) | 13,374 (44%) | 787 (46%) |
| Medium | 4,980 (16%) | 4,678 (15%) | 302 (18%) |
| High | 3,946 (12%) | 3,746 (12%) | 200 (12%) |
| Unknown | 1,432 | 1,385 | 47 |
| County, (%) |  |  |  |
| Iztapalapa | 18,428 (55%) | 17,379 (55%) | 1,049 (60%) |
| Coyoacan | 15,090 (45%) | 14,382 (45%) | 708 (40%) |
| *Lifestyle* | | | |
| Poor quality diet, (%) | 18,332 (55%) | 17,404 (55%) | 928 (53%) |
| Smoking, (%) |  |  |  |
| Never | 6,972 (21%) | 6,638 (21%) | 334 (19%) |
| Former | 9,470 (28%) | 8,791 (28%) | 679 (39%) |
| Current | 17,076 (51%) | 16,332 (51%) | 744 (42%) |
| Alcohol consumption, (%) |  |  |  |
| Never | 1,999 (6.0%) | 1,878 (5.9%) | 121 (6.9%) |
| Former | 3,107 (9.3%) | 2,981 (9.4%) | 126 (7.2%) |
| >3 times a month | 12,424 (37%) | 11,726 (37%) | 698 (40%) |
| >2 times a week | 15,988 (48%) | 15,176 (48%) | 812 (46%) |
| Physical activity, (%) |  |  |  |
| None | 22,988 (69%) | 21,740 (68%) | 1,248 (71%) |
| ≤2 times a week | 4,961 (15%) | 4,808 (15%) | 153 (8.7%) |
| ≥3 times a week | 5,569 (17%) | 5,213 (16%) | 356 (20%) |
| *Clinical features* | | | |
| Hypertension, (%) | 11,816 (35%) | 10,823 (34%) | 993 (57%) |
| Dyslipidemia, (%) | 29,494 (88%) | 27,984 (88%) | 1,510 (86%) |
| Overweight, (%) | 17,284 (52%) | 16,399 (52%) | 885 (50%) |
| Obesity, (%) | 8,229 (25%) | 7,771 (24%) | 458 (26%) |
| Metabolic syndrome, (%) | 8,996 (27%) | 8,387 (26%) | 609 (35%) |
| *Anthropometric* | | | |
| Body Mass Index, (kg/m2) | 27.4 (25.1, 29.9) | 27.4 (25.1, 29.9) | 27.5 (25.1, 30.1) |
| Waist circumference, (cm) | 95 (89, 101) | 95 (89, 101) | 97 (91, 103) |
| Waist-to-Height Ratio, (%) | 0.58 (0.54, 0.61) | 0.57 (0.54, 0.61) | 0.59 (0.56, 0.63) |
| *Biochemical assessment* | | | |
| Glucose, (mg/dl) | 72 (60, 86) | 72 (60, 85) | 77 (63, 91) |
| HbA1c, (%) | 5.45 (5.17, 5.63) | 5.45 (5.17, 5.63) | 5.54 (5.26, 5.81) |
| Total cholesterol, (mg/dl) | 161 (138, 184) | 161 (138, 184) | 158 (135, 181) |
| HDL-C, (mg/dl) | 36 (32, 40) | 36 (32, 40) | 35 (31, 40) |
| LDL-C, (mg/dl) | 65 (53, 77) | 65 (53, 77) | 63 (52, 75) |
| Triglycerides, (mg/dl) | 132 (102, 167) | 132 (102, 168) | 125 (98, 159) |
| *Surrogates evaluated* | | | |
| METSVF | 7.13 (6.85, 7.38) | 7.12 (6.84, 7.37) | 7.37 (7.13, 7.57) |
| CVAI | 135 (110, 162) | 134 (110, 161) | 151 (128, 177) |
| EVA | 127 (99, 157) | 125 (98, 155) | 152 (126, 180) |
| DAAT | 208 (172, 247) | 207 (171, 247) | 219 (182, 257) |
| DAI | 1.40 (1.08, 1.78) | 1.40 (1.08, 1.78) | 1.37 (1.05, 1.75) |
| VAI | 2.17 (1.66, 2.77) | 2.18 (1.66, 2.77) | 2.14 (1.63, 2.71) |
| LAAP | 44 (31, 61) | 44 (31, 61) | 45 (32, 62) |
| *Outcomes* |  |  |  |
| Cardiac-related deaths | 1,330 (4.0%) | 0 (0%) | 1,330 (76%) |
| Stroke-related deaths | 313 (0.9%) | 0 (0%) | 313 (18%) |
| Other-cause deaths | 114 (0.3%) | 0 (0%) | 114 (6%) |

Footnote: Variables are presented as median (interquartile range, IQR) or *n* (%).

Abbreviations: CVD, cardiovascular disease; HbA1c, glycosylated hemoglobin; HDL-C, HDL cholesterol; LDL-C, LDL cholesterol; METS-VF, Metabolic Score for Visceral Fat; CVAI, Chinese Visceral Adiposity Index; EVA, visceral adipose tissue area estimate; DAAT: deep abdominal adipose tissue; DAI: dysfunctional adiposity index; VAT: visceral adiposity index; LAP: lipid accumulation product

Supplementary Table 5. Baseline characteristics of female patients included from the Mexico City Prospective Cohort, stratified according to the presented outcome

| Characteristic | Overall  N = 68,867 | Outcome | |
| --- | --- | --- | --- |
|  |  | **Censored**  N = 66,556 | **Mortality due CVD**  N = 2,311 |
| *Demographics* | | | |
| Age, (years) | 46 (40, 56) | 46 (40, 55) | 66 (56, 73) |
| Social development index, (%) | | | |
| Very low | 19,696 (30%) | 19,103 (30%) | 593 (26%) |
| Low | 27,527 (42%) | 26,551 (42%) | 976 (44%) |
| Medium | 10,050 (15%) | 9,640 (15%) | 410 (18%) |
| High | 8,552 (13%) | 8,291 (13%) | 261 (12%) |
| Unknown | 3,042 | 2,971 | 71 |
| County, (%) |  |  |  |
| Iztapalapa | 41,481 (60%) | 40,031 (60%) | 1,450 (63%) |
| Coyoacan | 27,386 (40%) | 26,525 (40%) | 861 (37%) |
| *Lifestyle* | | | |
| Poor quality diet, (%) | 29,700 (43%) | 28,683 (43%) | 1,017 (44%) |
| Smoking, (%) |  |  |  |
| Never | 42,439 (62%) | 40,855 (61%) | 1,584 (69%) |
| Former | 9,597 (14%) | 9,230 (14%) | 367 (16%) |
| Current | 16,831 (24%) | 16,471 (25%) | 360 (16%) |
| Alcohol consumption, (%) |  |  |  |
| Never | 17,131 (25%) | 16,411 (25%) | 720 (31%) |
| Former | 3,243 (4.7%) | 3,170 (4.8%) | 73 (3.2%) |
| >3 times a month | 10,599 (15%) | 10,164 (15%) | 435 (19%) |
| >2 times a week | 37,894 (55%) | 36,811 (55%) | 1,083 (47%) |
| Physical activity, (%) |  |  |  |
| None | 55,647 (81%) | 53,759 (81%) | 1,888 (82%) |
| ≤2 times a week | 3,679 (5.3%) | 3,555 (5.3%) | 124 (5.4%) |
| ≥3 times a week | 9,541 (14%) | 9,242 (14%) | 299 (13%) |
| *Clinical features* | | | |
| Hypertension, (%) | 22,913 (33%) | 21,406 (32%) | 1,507 (65%) |
| Dyslipidemia, (%) | 65,389 (95%) | 63,202 (95%) | 2,187 (95%) |
| Overweight, (%) | 29,779 (43%) | 28,917 (43%) | 862 (37%) |
| Obesity, (%) | 26,612 (39%) | 25,564 (38%) | 1,048 (45%) |
| Metabolic syndrome, (%) | 29,378 (43%) | 27,861 (42%) | 1,517 (66%) |
| *Anthropometric* | | | |
| Body Mass Index, (kg/m2) | 28.7 (25.9, 31.8) | 28.7 (25.9, 31.8) | 29.4 (26.1, 32.9) |
| Waist circumference, (cm) | 91 (84, 98) | 91 (84, 98) | 96 (88, 103) |
| Waist-to-Height Ratio, (%) | 0.60 (0.55, 0.65) | 0.60 (0.55, 0.65) | 0.64 (0.59, 0.69) |
| *Biochemical assessment* | | | |
| Glucose, (mg/dl) | 74 (62, 86) | 73 (62, 86) | 79 (66, 94) |
| HbA1c, (%) | 5.45 (5.17, 5.72) | 5.45 (5.17, 5.72) | 5.63 (5.35, 5.81) |
| Total cholesterol, (mg/dl) | 167 (143, 191) | 167 (143, 190) | 169 (143, 193) |
| HDL-C, (mg/dl) | 40 (35, 45) | 40 (35, 45) | 39 (34, 45) |
| LDL-C, (mg/dl) | 65 (54, 77) | 65 (54, 77) | 65 (53, 78) |
| Triglycerides, (mg/dl) | 120 (93, 153) | 120 (93, 153) | 121 (95, 153) |
| *Surrogates evaluated* | | | |
| METSVF | 6.91 (6.59, 7.19) | 6.90 (6.58, 7.17) | 7.26 (7.01, 7.44) |
| CVAI | 114 (87, 141) | 112 (87, 140) | 149 (125, 172) |
| EVA | 109 (84, 135) | 108 (84, 134) | 142 (118, 164) |
| DAAT | 137 (106, 170) | 136 (106, 169) | 162 (130, 196) |
| DAI | 1.45 (1.09, 1.88) | 1.45 (1.09, 1.88) | 1.51 (1.15, 1.93) |
| VAI | 2.55 (1.91, 3.31) | 2.55 (1.91, 3.31) | 2.67 (2.02, 3.40) |
| LAAP | 45 (31, 63) | 45 (31, 62) | 51 (37, 68) |
| *Outcomes* |  |  |  |
| Cardiac-related deaths | 1,608 (2.3%) | 0 (0%) | 1,608 (70%) |
| Stroke-related deaths | 562 (0.8%) | 0 (0%) | 562 (24%) |
| Other-cause deaths | 141 (0.2%) | 0 (0%) | 141 (6.1%) |

Footnote: Variables are presented as median (interquartile range, IQR) or *n* (%).

Abbreviations: CVD, cardiovascular disease; HbA1c, glycosylated hemoglobin; HDL-C, HDL cholesterol; LDL-C, LDL cholesterol; METS-VF, Metabolic Score for Visceral Fat; CVAI, Chinese Visceral Adiposity Index; EVA, visceral adipose tissue area estimate; DAAT: deep abdominal adipose tissue; DAI: dysfunctional adiposity index; VAT: visceral adiposity index; LAP: lipid accumulation product.

Supplementary Table 6. Association and comparison of clinical surrogates that estimate visceral fat for cause-specific mortality

|  | Unadjusted | | | Adjusted | | |
| --- | --- | --- | --- | --- | --- | --- |
| **Surrogates** | **HR** | **95% CI** | **p-value** | **HR** | **95% CI** | **p- value** |
| **Cardiac-related deaths** | | | | | | |
| METS-VF | 2.83 | 2.69, 2.97 | <0.001 | 1.18 | 1.12, 1.25 | <0.001 |
| EVA | 2.25 | 2.17, 2.33 | <0.001 | 1.17 | 1.11, 1.22 | <0.001 |
| CVAI | 2.20 | 2.12, 2.29 | <0.001 | 1.16 | 1.11, 1.22 | <0.001 |
| DAAT | 1.54 | 1.49, 1.59 | <0.001 | 1.16 | 1.10, 1.21 | <0.001 |
| LAP | 1.16 | 1.12, 1.20 | <0.001 | 0.98 | 0.94, 1.02 | 0.4 |
| DAI | 1.02 | 0.98, 1.05 | 0.4 | 0.91 | 0.88, 0.95 | <0.001 |
| VAI | 0.98 | 0.95, 1.02 | 0.3 | 0.91 | 0.88, 0.95 | <0.001 |
| **Stroke-related deaths** | | | | | | |
| METS-VF | 2.57 | 2.35, 2.80 | <0.001 | 1.15 | 1.03, 1.27 | 0.010 |
| EVA | 2.05 | 1.92, 2.19 | <0.001 | 1.06 | 0.97, 1.16 | 0.2 |
| CVAI | 2.02 | 1.89, 2.16 | <0.001 | 1.03 | 0.94, 1.13 | 0.5 |
| DAAT | 1.37 | 1.28, 1.45 | <0.001 | 1.07 | 0.98, 1.17 | 0.15 |
| LAP | 1.15 | 1.08, 1.23 | <0.001 | 0.93 | 0.86, 1.00 | 0.062 |
| DAI | 1.05 | 0.98, 1.12 | 0.2 | 0.91 | 0.85, 0.98 | 0.012 |
| VAI | 1.04 | 0.98, 1.11 | 0.2 | 0.91 | 0.85, 0.98 | 0.019 |
| **Other-cause deaths** | | | | | | |
| METS-VF | 2.75 | 2.64, 2.87 | <0.001 | 1.11 | 0.92, 1.34 | 0.3 |
| EVA | 2.20 | 1.95, 2.47 | <0.001 | 1.18 | 1.00, 1.39 | 0.055 |
| CVAI | 2.14 | 1.89, 2.43 | <0.001 | 1.15 | 0.97, 1.37 | 0.10 |
| DAAT | 1.52 | 1.36, 1.71 | <0.001 | 1.12 | 0.96, 1.32 | 0.2 |
| LAP | 1.21 | 1.08, 1.36 | 0.001 | 1.02 | 0.88, 1.17 | 0.8 |
| DAI | 1.10 | 0.98, 1.24 | 0.11 | 0.98 | 0.86, 1.13 | 0.8 |
| VAI | 1.06 | 0.94, 1.20 | 0.3 | 0.99 | 0.86, 1.14 | 0.9 |

Footnote: Models were adjusted for age, sex, poor-quality diet and metabolic syndrome.

Abbreviations: HR, hazard ratio; CI, confidence interval.

Supplementary Table 7. Comparison of the performance of the Globorisk model with and without the addition of METS-VF across overall and different risk strata

| Risk groups | GloboRisk  (C-Statistic, 95% CI) | GloboRisk / METSVF  (C-Statistic, 95% CI) | Δ C Statistic | Δ BIC |
| --- | --- | --- | --- | --- |
| All | 0.781 (0.78-0.79) | 0.784 (0.78-0.79) | 0.003 | 3,606.62 |
| Very Low Risk | 0.68 (0.66-0.69) | 0.69 (0.68-0.7) | 0.016 | 736.18 |
| Low Risk | 0.55 (0.53-0.56) | 0.56 (0.54-0.57) | 0.008 | 631.93 |
| Intermediate Risk | 0.52 (0.5-0.54) | 0.56 (0.54-0.58) | 0.042 | 169.12 |
| High Risk | 0.53 (0.52-0.55) | 0.54 (0.52-0.56) | 0.009 | 361.26 |
| Very High Risk | 0.55 (0.54-0.57) | 0.56 (0.55-0.57) | 0.008 | 1,114.58 |

Footnotes:

- Risk groups were stratified according to the GloboRisk index in very low risk (< 3), low risk (≥ 3 & < 7), intermediate risk (≥ 7 & < 10), high risk (≥ 10 & < 15), and very high risk (≥ 15).
- ΔC statistic were obtained by the subtraction of the c statistics of Globo Risk minus GloboRiks/METS-VF, and it shown on natural numbers
- Δ BIC were obtained by the subtraction of BIC values of GloboRisk minus GloboRisk/METS-VF.

Abbreviations: BIC, Bayesian Information Criterion.
